# Clustering schizophrenia genes by their temporal expression patterns aids functional interpretation *genetics-based evidence in favor of the two-hit hypothesis*

**DOI:** 10.1101/2022.08.25.22279215

**Authors:** Dennis van der Meer, Weiqiu Cheng, Jaroslav Rokicki, Sara Fernandez-Cabello, Alexey Shadrin, Olav B. Smeland, Friederike Ehrhart, Sinan Gülöksüz, Nils Eiel Steen, Srdjan Djurovic, Lars T. Westlye, Ole A. Andreassen, Tobias Kaufmann

**Affiliations:** Norwegian Centre for Mental Disorders Research (NORMENT), Division of Mental Health and Addiction, Oslo University Hospital & Institute of Clinical Medicine, University of Oslo, Oslo, Norway; School of Mental Health and Neuroscience, Faculty of Health, Medicine and Life Sciences, Maastricht University, The Netherlands; Department of Psychology, University of Oslo, Oslo, Norway; Department of Psychiatry, Yale University School of Medicine, New Haven, Connecticut; Department of Medical Genetics, Oslo University Hospital, Oslo, Norway; Norwegian Centre for Mental Disorders Research, Department of Clinical Science, University of Bergen, Bergen, Norway; K.G. Jebsen Centre for Neurodevelopmental Disorders, University of Oslo; Department of Psychiatry and Psychotherapy, Tübingen Center for Mental Health, University of Tübingen, Germany

**Keywords:** schizophrenia, gene expression, two-hit hypothesis, neurodevelopment

## Abstract

Schizophrenia is a highly heritable brain disorder with a typical symptom onset in early adulthood. The two-hit hypothesis posits that schizophrenia results from deviant early neurodevelopment, predisposing an individual, followed by a disruption of later brain maturational processes that trigger the onset of symptoms. Here, we investigate how the timing of expression of 345 putative schizophrenia risk genes may aid in understanding the interplay of neurobiological processes in the pathophysiology of schizophrenia. Clustering of brain transcriptomic data across the lifespan revealed a set of 183 genes that was significantly upregulated prenatally and downregulated postnatally and 162 genes that showed the opposite pattern. The prenatally upregulated set of genes was functionally annotated to fundamental cell cycle processes, while the postnatally upregulated set was associated with the immune system and neuronal communication. We subsequently calculated two set-specific polygenic risk scores for 743 individuals with schizophrenia and 743 sex- and age-matched healthy controls. We found an interaction between the two scores; higher prenatal polygenic risk was only significantly associated with schizophrenia diagnosis and severity, at higher levels of postnatal polygenic risk. We therefore provide genetics-based evidence in favor of the two-hit hypothesis, supporting that schizophrenia may be shaped by disruptions of separable biological processes acting at distinct phases of neurodevelopment.

---

Schizophrenia is a severe brain disorder that is highly heritable,^1–3^ determined by a large number of interacting genetic and environmental factors.^4^ Its clinical onset is typically in early adulthood, yet there are behavioral and biological indicators present many years before this^5^, with deviations of neurodevelopmental trajectories early in life.^3^

Schizophrenia is a dynamic and heterogenous disorder, and the early development and clinical course of the illness reflects many biological processes interacting over time. The classic two-hit hypothesis of schizophrenia posits that the clinical phenotype stems from a combination of early-acting risk factors that predispose an individual, followed by a second “hit” at a later stage of development that leads to the onset of symptoms and subsequent diagnosis.^6^ The first, priming hit is thought to disrupt neurogenesis and differentiation in early neurodevelopmental phases, while the second hit may involve processes more related to neuroplasticity.^7^ Inflammatory processes have been implicated in this chain of events, with several lines of evidence indicating a link between the immune system and the development of schizophrenia,^8^ albeit an enigmatic one.^9^ Given the complex etiology and heterogeneous clinical manifestation of schizophrenia, the two-hit hypothesis is undoubtedly an oversimplification that bins numerous biological processes and their waxing and waning effects over time. Yet, the underlying notion that risk factors act primarily at different stages of development, moderating each other’s influence on neurodevelopmental trajectories, represents an often-overlooked developmental dynamic dimension, which can explain some of the observed etiological and clinical heterogeneity of schizophrenia.

Genome-wide association studies (GWAS) of schizophrenia have discovered hundreds of associated common genetic variants, mapped onto genes expressed primarily in the brain, and linked to pathways involved in immune system regulation and synaptic functioning.^10,11^ The effects of genetic variants on neurodevelopment are likely to be age-dependent,^12^ as also suggested by strong changes in gene expression levels over the lifespan.^13^ Further, genes that are expressed together are more likely to participate in the same functional processes.^14^ This indicates that the temporal expression patterns of genes or gene-sets reflect biological processes relevant for the risk and clinical course of schizophrenia at distinct phases of disease development. Here, we clustered genes previously found to be significantly associated with schizophrenia into two groups based on their age-associated expression patterns, allowing us to infer dynamic processes involved in schizophrenia.

## Results

First, we sought to identify genes associated with schizophrenia, regardless of the timing of their expression. To this end, we used gene-based tests,^15^ aggregating effects of variants across each gene, applied to the European-specific Psychiatric Genomics Consortium schizophrenia GWAS wave 3 summary statistics.^10^ Out of the 508 significant genes identified through this procedure, 345 protein-coding genes had brain expression data available from 55 clinically unremarkable donors ranging from 56 days post-conception up to 82 years in age.^13,16^ The mean expression across these 345 genes and over all cortical brain samples was highest prenatally, in line with the framing of schizophrenia as a neurodevelopmental disorder.^3,17^ For comparison, we also calculated the mean expression across 82 genes associated with Alzheimer’s disease,^18^ i.e., a neurodegenerative disease affecting older (>65yrs) people, as well as across all 16,660 genes with available expression data, both of which showed a distinct pattern with higher peaks of expression postnatally (**Figure 1a**).

**Figure 1.**
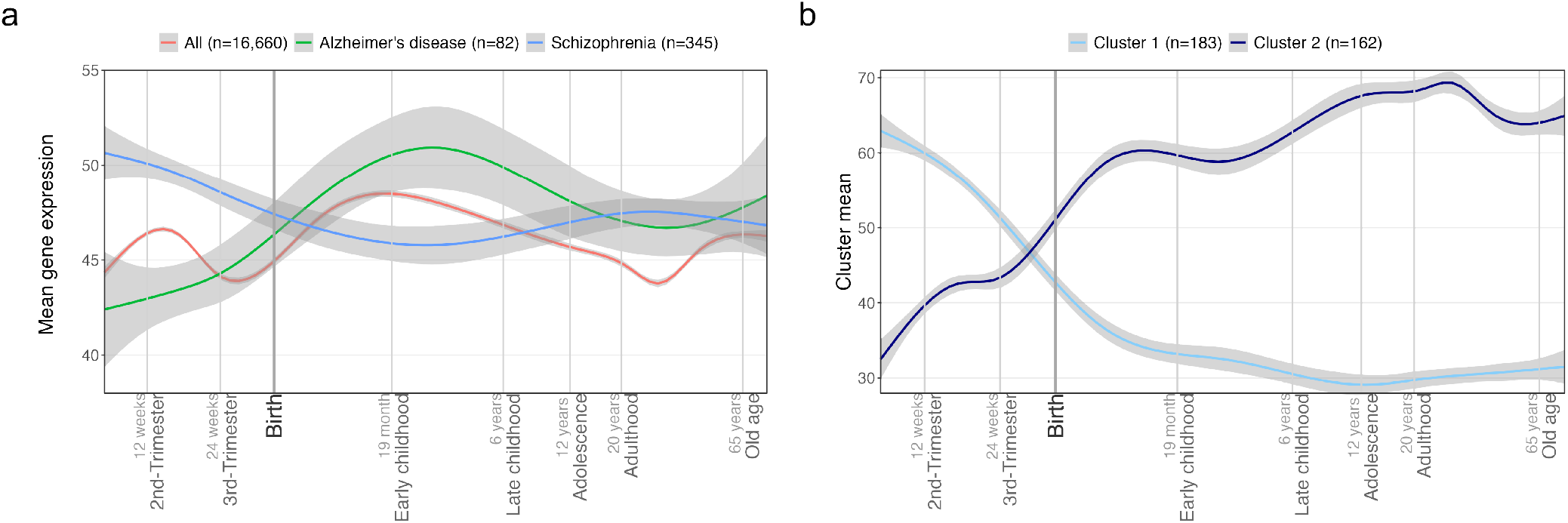
Mean gene expression over the lifespan. a) Mean expression (y-axis) over time (x-axis) for genes significantly associated with schizophrenia (SCZ, in blue), Alzheimer’s disease (AD, in red), or all genes (green). b) mean expression over time, for each of the two sets of genes (shades of blue-coloured lines) identified through hierarchical clustering applied to the expression of the schizophrenia genes. Lines were fitted through generalized additive modelling, with the grey shading reflecting the standard error 0.95 confidence interval.

We applied hierarchical clustering to the data, grouping the putative schizophrenia genes by similarity of the scaled expression patterns over time, through Ward clustering^19^ based on Euclidian distances. We selected a two-cluster solution, in line with the two-hit hypothesis and as deemed optimal by clustering indices (**Supplementary Figure 1**; the list of genes per cluster is provided in **Supplementary Table 1)**. The distinction between the two clusters of genes, based on their mean expression over time, can be most clearly characterized as being predominantly expressed prenatally (peak early in pregnancy) versus postnatally (peak in adulthood) (**Figure 1b**). Analyses of differential expression of the genes through the FUMA pipeline^20^ recapitulated these findings, indicating highly significant (minimum p=8.4×10^−10^) upregulation in prenatal brain tissue and downregulation postnatally for the first cluster, and vice versa (minimum p=1.1×10^−11^) for the second cluster, see **Supplementary Figure 2**. Hence, we will refer to these as the pre- and the postnatal cluster.

The two clusters were associated with distinct biological processes, as indicated by functional annotation based both on Reactome^21^ and Gene Ontology.^22^ On the highest level of hierarchy of Reactome, the prenatal cluster was significantly associated with chromatin organization (p=5.0×10^−7^), reproduction (p=3.3×10^−6^), DNA replication (6.0×10^−5^), DNA repair (p=2.4×10^−4^), cell cycle (p=3.2×10^−4^), and cellular responses to stimuli (7.7×10^−4^). The postnatal cluster on the other hand was associated with the immune system (p=2.3×10^−4^), disease (p=2.7×10^−4^) and the neuronal system (p=4.6×10^−4^). This is visualized in **Supplementary Figure 3**, with the full list of enriched terms at lower levels of the hierarchy provided in **Supplementary Table 2**. In agreement with this, the most significant enrichment among 22,749 individual Gene Ontology processes were ‘regulation of DNA-templated transcription’ (p=1.2×10^−8^) for the prenatal cluster and ‘neuron projection’ (p=5.2×10^−12^) for the postnatal cluster. In order to check whether the clustering truly aided functional interpretation, we randomly selected 5000 subsets of genes of the same size as the two clusters (n=183 and n=162) from either all protein-coding genes (N=19,427) or from the set of schizophrenia risk genes (N=345). We then performed the same gene-set enrichment analyses on each of these random sets and recorded the smallest observed p-value for each of the 5000 runs. The results, displayed in **Figure 2**, indicated that the enrichment reached for the pre- and postnatal cluster were far above chance level in either case. Therefore, clustering genes based on their age-associated expression patterns aided in identifying distinct biological processes contributing to schizophrenia.

**Figure 2.**
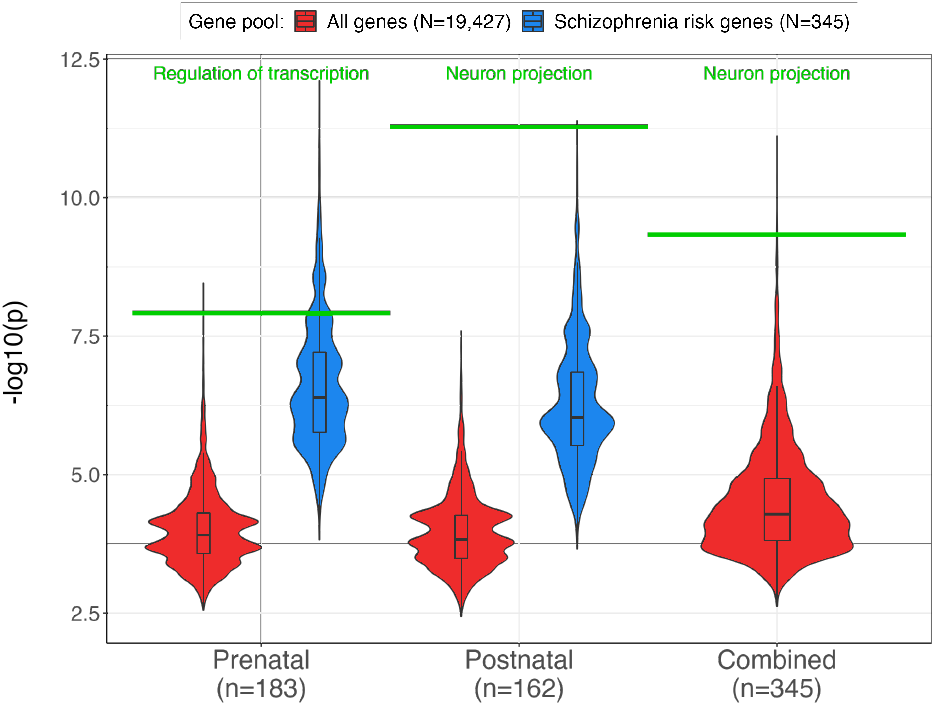
Functional annotation of the two gene clusters. Significance (y-axis) of the most significant Gene Ontology pathway for each of 5000 randomly drawn sets of genes of the same size as either the prenatal, postnatal, or both combined (x-axis). Red violins show the -log10(p-value) distribution for random gene sets when drawn out of the entirety of protein-coding genes, and blue violins show this distribution for random gene sets when drawn out of the smaller pool of schizophrenia genes. The green horizontal line indicates the -log10(p-value) of the most significant pathway, listed at the top in green, for the true pre- and postnatal gene sets.

Using gene-based analyses through MAGMA, applied to the latest GWAS summary statistics, we checked the association of the two gene clusters with other brain disorders. As shown in Figure 3, these clusters were significantly associated with several brain disorders, albeit to differing degrees. Most notable is that major depressive disorder (MDD) was only highly significantly associated with the postnatal cluster, not the prenatal, suggesting that its genetic overlap with schizophrenia^23,24^ is mostly driven by genes expressed postnatally. In contrast, the data from the autism spectrum disorder (ASD) GWAS revealed the opposite pattern, i.e., ASD was associated with the prenatal gene cluster but not the postnatal cluster, in line with its early onset.

**Figure 3.**
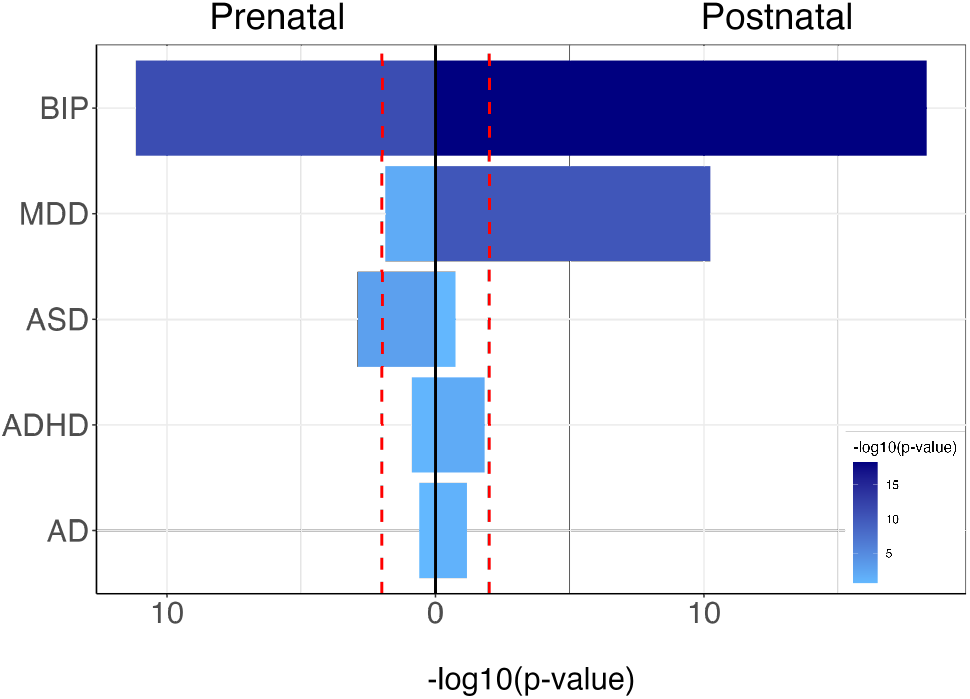
Association of brain disorders (y-axis) with the sets of schizophrenia genes expressed prenatally (left) or postnatally (right), with the x-axis indicating significance in -log10(p-value) of the gene-based analyses. The fill color also reflects significance, as shown in the legend, with the vertical red dotted lines indicating the Bonferroni significance threshold at p=.01. Abbreviations: BIP=bipolar disorder, MDD=Major depressive disorder, ASD=Autism spectrum disorder, ADHD=Attention-deficit hyperactivity disorder, AD=Alzheimer’s disease.

Next, we constructed cluster-specific schizophrenia polygenic scores by clumping only variants within genes belonging to either cluster, using PRSet^25^. We tested for associations with both cluster scores in data from 743 White European individuals with schizophrenia and 743 age- and sex-matched healthy controls using logistic regression, with diagnosis as outcome measure and the two mean-centered polygenic scores, as well as mean-centered age, sex and twenty genetic principal components as the predictors. We included an interaction term between the two cluster-specific polygenic scores to test the two-hit hypothesis, that the occurrence of schizophrenia can be explained by the joint effect of two temporally distinct biological processes. We found that there was indeed a significant interaction effect (β=.11, p=.02), as visualized in **Figure 4**, such that the association of the prenatal cluster score (conditional β=.16, p=.004) with schizophrenia diagnosis was positively moderated by the postnatal score (conditional β=.17, p=.003). The full output of the model is listed in Supplementary Table 3.

**Figure 4.**
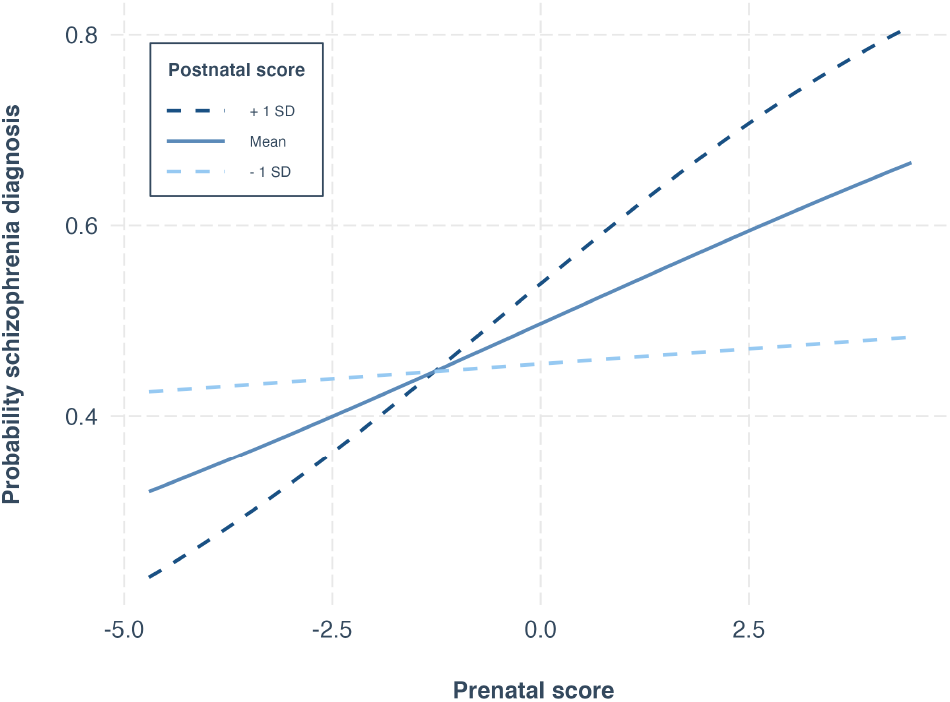
Association between the prenatal polygenic score (x-axis) and schizophrenia diagnosis (y-axis), moderated by the postnatal polygenic score (line type and colour). SD=standard deviation.

## Discussion

The two-hit hypothesis posits that the risk and clinical course of schizophrenia are dependent on the joint effects of temporally distinct biological processes occurring during different phases of development. Here, we clustered genes associated with schizophrenia based on their age-dependent expression patterns, and showed that the genes belonging to the resulting two clusters are transcribed predominantly prenatally versus postnatally. Through functional annotation, we discovered that such a division contributes to characterization of the biological processes involved in schizophrenia. We additionally found that variation in postnatally expressed genes moderated the effects of variation in prenatally expressed genes on schizophrenia, illustrating the importance of integrating temporal dynamics of schizophrenia into genetic research.

We found that putative schizophrenia risk genes implicated by common genetic variants are expressed above average early in life, in line with the neurodevelopmental conceptualization of the disorder,^3,17^ and corroborating findings of other gene expression studies.^26^ However, our clustering approach further divided schizophrenia genes into sets that are predominantly expressed prenatally or postnatally. Importantly, for both sets, functional annotation reached greater significance than when taking random subsets of genes of the same size from the total set of schizophrenia genes. This shows that the division based on estimated age-associated expression patterns succeeded in capturing distinct processes. The prenatally expressed set of genes was functionally annotated to fundamental cell cycle processes. Fitting the hypothesized first hit, perturbations of such early processes are likely to affect neurodevelopmental trajectories, increasing the likelihood of dysfunctional neural circuits that mature in adolescence, which may underly the onset of symptoms.^7^ Annotation of the postnatally expressed set of genes indicated they are primarily involved in the adaptive immune system and neuronal communication. The identification of components of the immune system, often linked to schizophrenia etiology, may be due to their important role in synaptic pruning, taking place during childhood and adolescence.^9,27^ This would fit with the identification of other significant pathways that are in line with the longstanding characterization of schizophrenia as a disorder of synaptic functioning and dysconnectivity.^10,28^

Comorbidity of schizophrenia with a range of other brain disorders is likely to be partly due to the central role of the biological processes captured by the two sets of genes in neurodevelopment. While bipolar disorder was strongly associated with both sets, ASD and MDD were only associated with the prenatal or postnatal cluster, which may be informative with regard to characterizing their comorbidity with schizophrenia. The observed pattern of associations of these gene sets with brain disorders that vary in their onset is thereby in line with the notion that schizophrenia results from a combination of dysfunction in both early and later in life brain maturation processes.

By combining information from GWAS with expression data from clinically unremarkable donors across the lifespan, this study complements work focusing on genes with differential expression between cases and controls,^29^ as those may overlook the effects of genes acting early, before diagnosis, and be confounded by secondary disease processes. Indeed, previous work has found that gene expression differs between individuals in different clinical stages of schizophrenia.^30^ Our results are thereby in accordance with the two-hit hypothesis of schizophrenia, that deviant brain maturation exacerbates earlier insults to the system to bring about the disorder. We provide genetics-based evidence of this hypothesis, captured by an interaction term between the set-specific polygenic scores; while the ‘hits’ are generally assumed to be environmental due to the temporal dimension, they may also be partly genetic, considering that genes exert their effects at specific times depending on when they are expressed. The amount of expression, and its impact on molecular processes downstream, can thereby be moderated both by environmental influences and genetic variation. The set-specific polygenic scores used in this study thereby provide a way to aggregate weak individual genetic effects acting on specific pathways and at specific time points, which enable us to study their role in neurodevelopment.

Overall, this study provides a proof-of-concept that information on the timing of gene expression aids the modelling of genetic effects on schizophrenia, albeit with several notable limitations. Given the complexity of schizophrenia and the continuous nature of the biological processes underlying it, the two-hit hypothesis is thereby less likely to adequately describe the etiology of schizophrenia than a multi-hit model or a continuum.^31^ It should be noted that detection of (higher-order) interactions requires a substantial amount of statistical power,^32^ and that there are known issues with modeling of epistasis for individual variants.^33^ Replication of these findings will therefore require well-powered samples, and it will be of interest to investigate potential clinical and demographic modifiers. Future studies could further look into the moderating effects of exposure to environmental factors during specific phases, investigation of specific symptom domains or other clinical characteristics as outcome measures, and brain regional specificity of the identified effects. The approach used to map genetic variation to genes may further be developed; in the current context, gene-based tests offer advantages over locus mapping by aggregating more information. Information on eQTLs and chromatin interactions may further improve the identification of the genes involved, yet currently available annotations for common gene-based tests are specific to a developmental phase, which would bias our results.

To conclude, the modelling of genetic risk factors that moderate each other’s effect, informed by the timing of their expression or occurrence, will aid in a better understanding of the development of schizophrenia.

## Data Availability

The data incorporated in this work were gathered from public resources. The code is available via https://github.com/norment/open-science [published upon acceptance]. Correspondence and requests for materials should be addressed to

## Methods

All data processing and statistical analyses were carried out through R v4.1.0,^34^ unless specified otherwise, with code available via https://github.com/norment/open-science.

### Gene expression data processing

We made use of gene expression data derived from brain tissue from 55 clinically unremarkable donors ranging in age from 8 weeks post conception to 82 years.^13^ We took the data as preprocessed by Kang *et al*., selecting for each gene the probe with the highest differential stability, n=16,660. Given the relatively high homogeneity of expression patterns across cortical brain samples,^35^ we subsequently averaged over 13 cortical regions, within donor, and scaled the expression values, within probe, across donors, to a range between 0 (lowest observed value) and 100 (highest observed value).

### Gene-based analyses

We used the summary statistics from the Psychiatric Genomics Consortium, wave 3 (PGC3) schizophrenia GWAS^10^ to identify schizophrenia risk genes, restricted to the European cohorts. We selected a version of the meta-analyzed summary statistics excluding the TOP sample, preventing sample overlap. This version contained 13,025,668 SNPs for 50,965 individuals with schizophrenia and 68,049 controls. For Alzheimer’s disease,^18^ attention deficit hyperactivity disorder,^36^ MDD,^37^ ASD,^38^ and bipolar disorder^39^ we made use of the latest GWAS summary statistics, as described in the references. For bipolar disorder we also ensured to use data with the TOP sample excluded.

We carried out gene-based tests using MAGMA v1.08 with default settings, which entails the application of a SNP-wide mean model and use of the 1000 Genomes Phase 3 EUR reference panel. For schizophrenia, 508 genes had a p-value smaller than 0.05/19,047, surviving multiple comparisons correction. Of these, 431 had a probe in the brain expression dataset, and of those 345 were protein-coding genes with evidence of being expressed in the brain, as summarized by the human protein atlas project (www.proteinatlas.org).^40^ We chose gene-based tests over locus mapping as the downstream analyses were all gene-rather than variant-centric, with these tests thereby aggregating a greater amount of information for the purpose of identifying the most relevant genes. Integration of information on eQTLs and chromatin interactions may allow for further improvements in the identification of genes involved based on GWAS data. However, recently introduced tools that enable this, such as e-MAGMA^41^ or H-MAGMA,^42^ work with annotation files based on e.g. fetal or adult tissue. This age-specificity precludes their use in this study, as they would bias the results.

Functional annotation of the clusters was achieved by uploading the gene lists to Reactome (https://reactome.org/), and running the pathway analysis with default settings. A list of all enriched pathways, significant after multiple comparisons correction through false discovery rate (FDR), is provided in Supplementary Table 1. Gene-set enrichment analyses were further carried out with the *topGO* function, part of the limma R package, checking for enrichment among 22,749 Gene Ontology processes, as listed in the Molecular Signatures Database (MsigdB; v7.1).

Plotting of the mean expression over time per gene set was done with *ggplot2* in R v4.0.3., with geom_smooth(method=“gam”) using default settings.

### Clustering of the expression data

We first checked for the optimal number of components among the scaled expression data of the schizophrenia gene set, using the NbClust R package with distance set to Euclidean and method set to ward.D2.^19^ Across a range of indices, there was most support for a two component solution (7), followed by a four (6) and three (5) component solution, see Supplementary Figure 1. In line with our aim to test the two-hit hypothesis of schizophrenia and in accordance with the majority rule, we selected the two-component solution. We then applied hierarchical clustering, through the *hclust* function, to the Euclidian distance matrix of this expression data, with the ward.D2 method.

### Polygenic score analyses

We selected data from White European participants of the Thematically Organised Psychosis (TOP) clinical cohort for the polygenic scoring analyses. We had complete genetic and covariate data available for 743 individuals with schizophrenia (mean age 32.76 years, SD=13.29; 42.80% female) and 1074 healthy individuals. The healthy individuals were then matched to those with schizophrenia on age and sex, using default settings of the MatchIt package, keeping 743 healthy individuals (mean age 31.66 years, SD=11.04; 42.66% female). White European ethnicity was based on self-report. DSM-IV diagnosis of schizophrenia was determined based on the Structured Clinical Interview for DSM-IV Axis I Disorders (SCID), carried out by trained physicians or clinical psychologists. Controls were individuals without brain damage or a lifetime history of a severe psychiatric disorder themselves or in first-degree relatives. Each sample was collected with the participants’ written informed consent and with approval by local Institutional Review Boards.

We used the PRSet functionality of PRSice-2 to make set-specific polygenic scores.^25,43^ These scores were based on the effect sizes of lead SNPs within the genomic boundaries of the genes that made up the two sets, with these boundaries being based on genome build GTCh37.

To analyze the association between the set-specific polygenic scores and schizophrenia, we used logistic regression, with diagnosis as outcome measure and the two polygenic scores, as well as age, sex and twenty genetic principal components as the predictors. We also included an interaction term between the two cluster-specific polygenic scores. See Supplementary Table 3 for the full model.

## Acknowledgements

The authors were funded by the Research Council of Norway (276082, 323961, 213837, 223273, 204966/F20, 229129, 249795/F20, 225989, 248778, 249795, 298646, 300767), the South-Eastern Norway Regional Health Authority (2013-123, 2014-097, 2015-073, 2016-064, 2017-004, 2019-101, 2019-107, 2020-086), Stiftelsen Kristian Gerhard Jebsen (SKGJ-Med-008), The European Research Council (ERC) under the European Union’s Horizon 2020 research and innovation programme (ERC Starting Grant, Grant agreement No. 802998).

This work was partly performed on the TSD (Tjeneste for Sensitive Data) facilities, owned by the University of Oslo, operated and developed by the TSD service group at the University of Oslo, IT-Department (USIT). We thank the PGC working groups of AD, ADHD, ASD, BIP, MDD and SCZ for providing GWAS summary statistics.

## Author contributions

D.v.d.M. and T.K. conceived the study; D.v.d.M., J.R. and T.K. pre-processed the data. D.v.d.M. performed all analyses, with conceptual input from T.K.; All authors contributed to interpretation of results; D.v.d.M. drafted the manuscript and all authors contributed to and approved the final manuscript.

## Competing interests

Dr. Andreassen has received speaker’s honorarium from Lundbeck, and is a consultant to HealthLytix. The other authors declare no competing financial interests.

## Notes

### Competing Interest Statement

Dr. Andreassen has received speaker honorarium from Lundbeck, and is a consultant to HealthLytix. The other authors declare no competing financial interests.

### Author Declarations

The Oslo Regional Committee for Medical Research Ethics and the Norwegian Data Inspectorate gave ethical approval for this study.

