## Supplementary Information for "Clustering schizophrenia genes by their temporal expression patterns aids functional interpretation *genetics-based evidence in favor of the two-hit hypothesis*"

**Supplementary Figures**

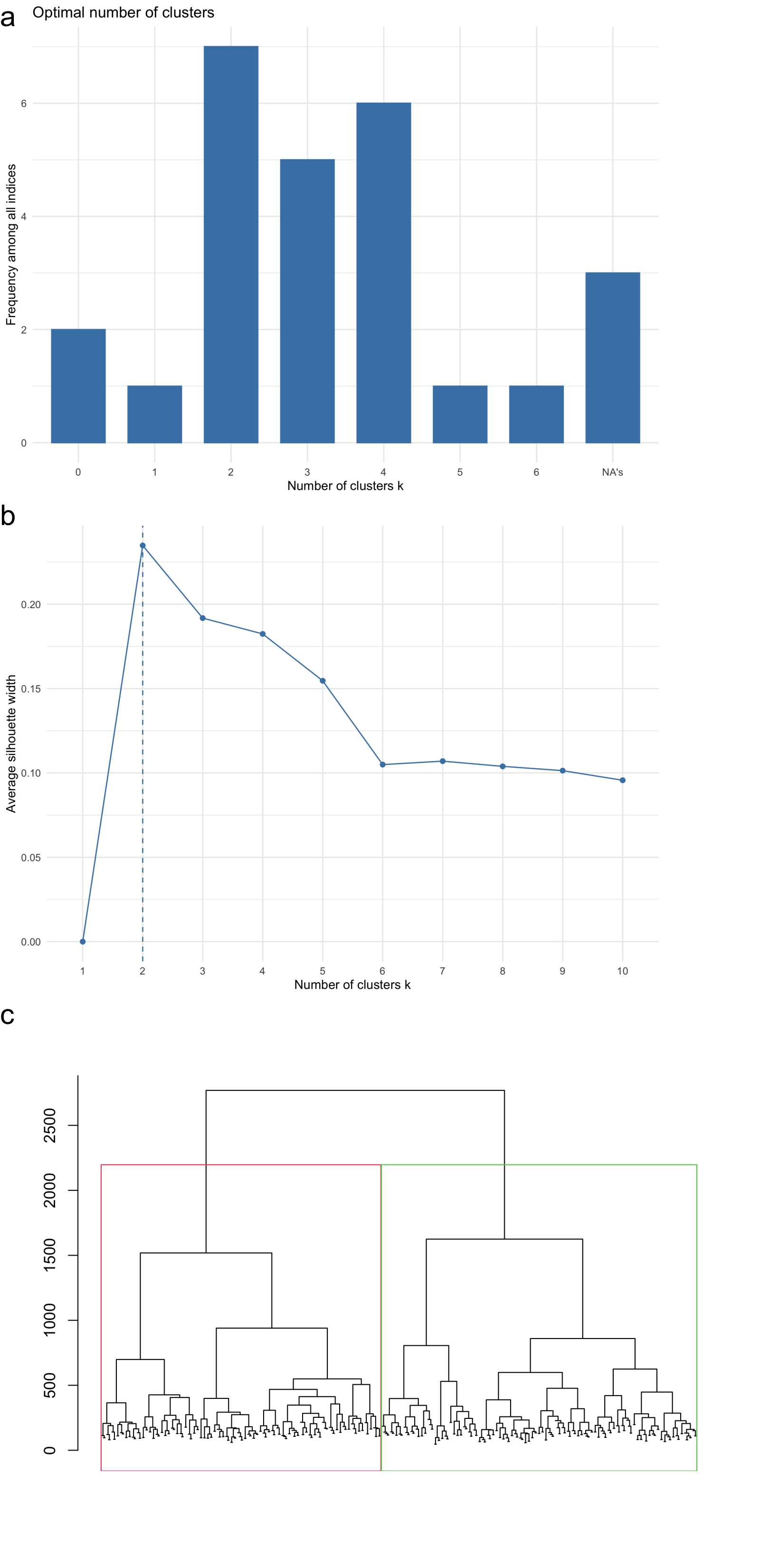

***Supplementary Figure 1. Selection of number of components for clustering the age-associated gene expression data****. a) A bar plot listing the number of clustering indices (y-axis) that indicated the number of components on the x-axis was the optimal solution. According to the majority rule, two components is optimal. b) Chart showing on the y-axis the average silhouette width, a common metric of how well the data can be clustered, for each of number of components, shown on the x-axis.*  *c) Dendrogram for the hierarchical clustering of genes based on their expression over the lifespan. The colored boxes indicate the division of leaves into the two separate clusters.*

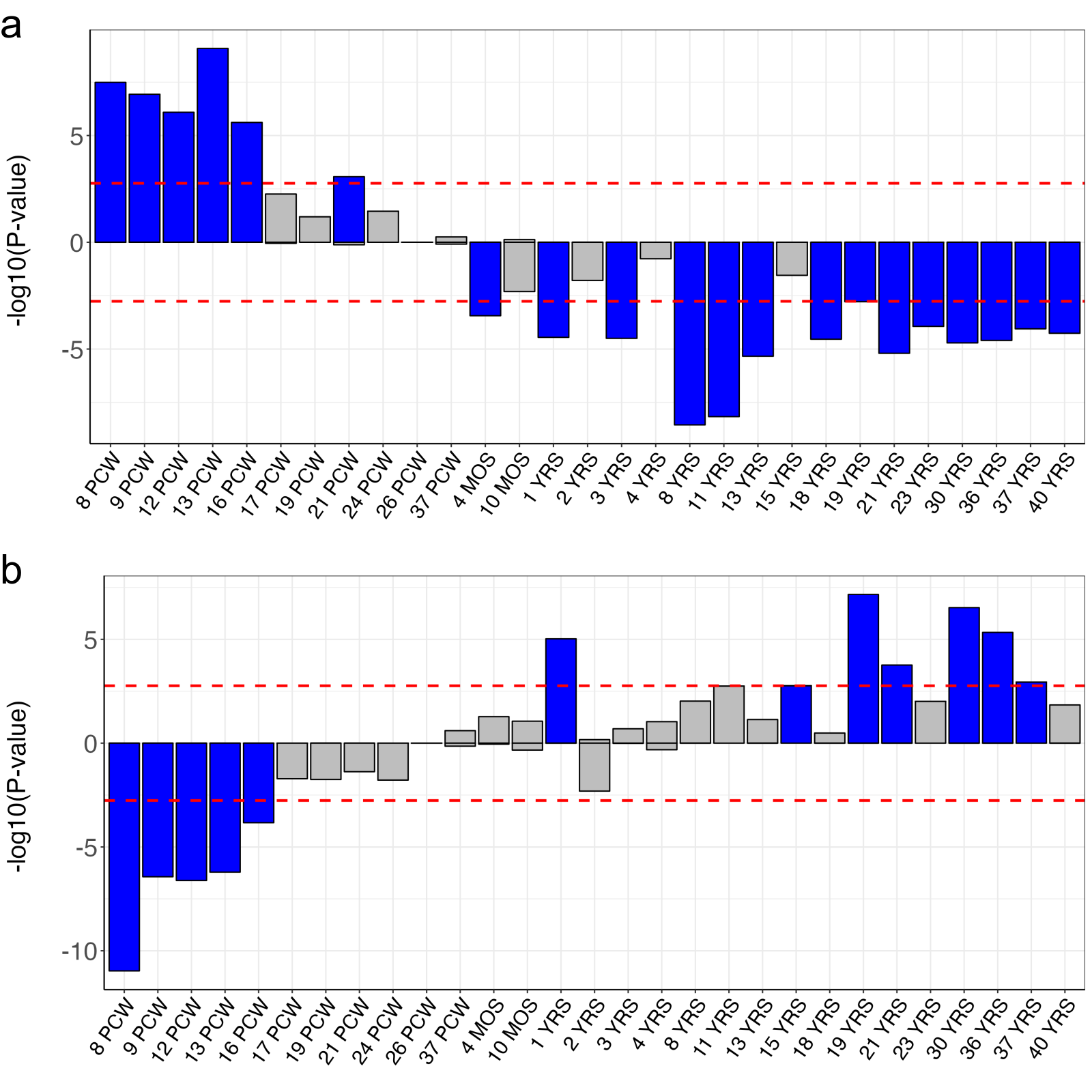

***Supplementary Figure 2.*** *Bar plots showing the significance (as -log10(p-value), on the y-axis) of differential expression tests of the two gene sets in the BrainSpan dataset, as available through FUMA, per age category (on the x-axis). The horizontal dotted line indicates the threshold for Bonferroni corrected significance, while the blue fill indicates that a bar has passed this threshold. Plot a) at the top shows the results for the prenatal set, and b) at the bottom for the postnatal set. PCW=post-conception weeks. MOS=months, YRS=years.*

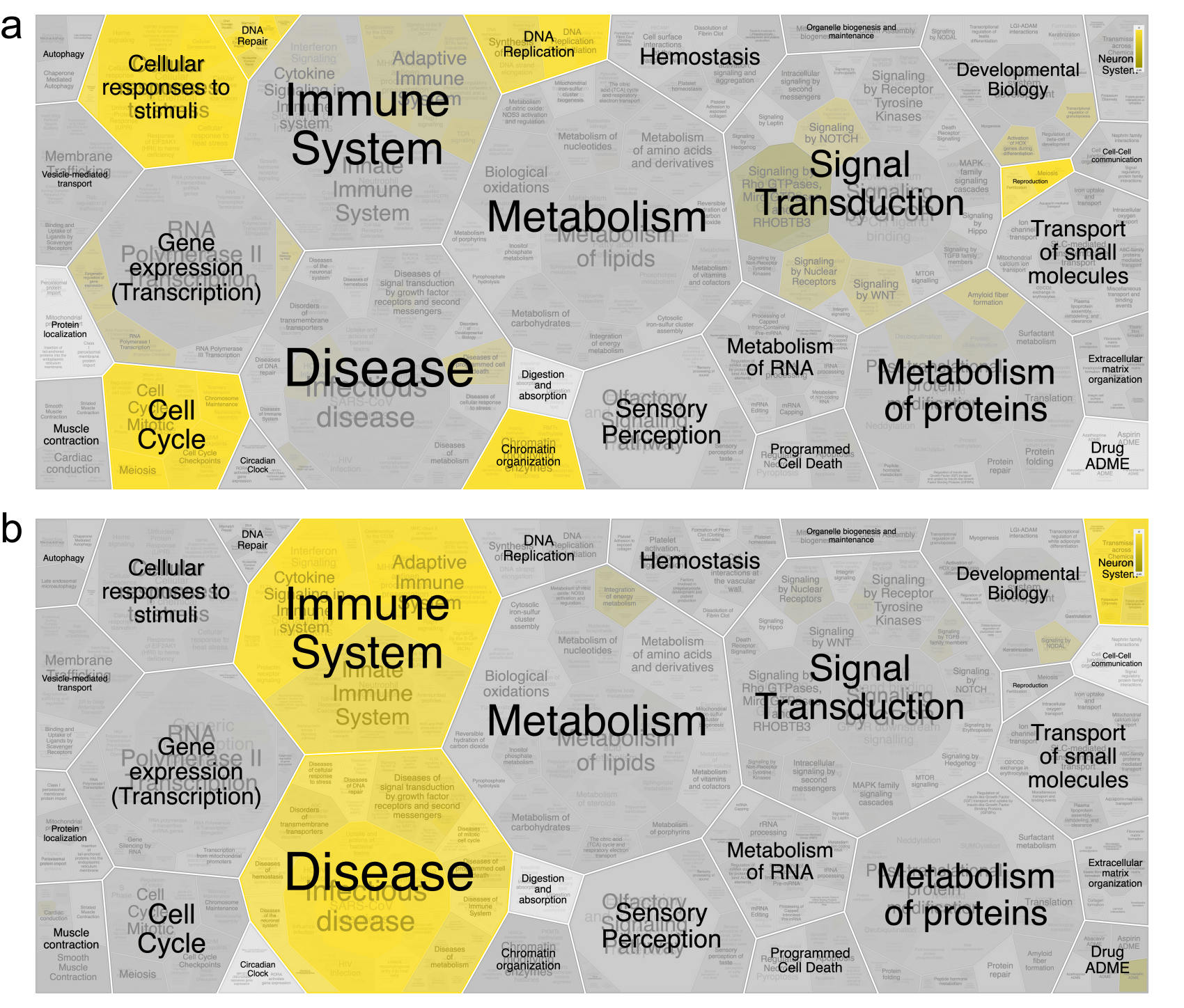
***Supplementary Figure 3.*** *a) Output from coupling the list of prenatally expressed genes to the Reactome database. Here areas highlighted in yellow indicate biological processes that are significantly enriched, while grey areas indicate lack of significance. b) same as for a) but for the postnatally expressed set of genes. For both a) and b) the significance of the processes is provided in the Results section of the manuscript, and in Supplementary Table 2.*

**Supplementary Tables**

Supplementary Tables 1 and 2 are provided as separate tab-delimited text files due to their length.

***Supplementary Table 3. Output of the full logistic regression model testing the association between the polygenic scores and schizophrenia diagnosis.***

| *Term* | *Beta* | *SE* | *P-value* |
| --- | --- | --- | --- |
| Intercept | -0.03 | 0.08 | 0.70 |
| Prenatal score | 0.16 | 0.06 | 4.4e-03 |
| Postnatal score | 0.17 | 0.06 | 2.6e-03 |
| Interaction | 0.13 | 0.06 | 0.02 |
| Age | 0.1 | 0.05 | 0.07 |
| Sex | 0.03 | 0.11 | 0.76 |
| Genetic PC1 | 0.09 | 0.06 | 0.15 |
| Genetic PC2 | 0.21 | 0.08 | 7.4e-03 |
| Genetic PC3 | -0.11 | 0.08 | 0.17 |
| Genetic PC4 | -0.01 | 0.07 | 0.88 |
| Genetic PC5 | -0.06 | 0.06 | 0.30 |
| Genetic PC6 | 0.17 | 0.07 | 0.01 |
| Genetic PC7 | -0.03 | 0.06 | 0.61 |
| Genetic PC8 | 0.18 | 0.06 | 2.2e-03 |
| Genetic PC9 | -0.01 | 0.06 | 0.90 |
| Genetic PC10 | 0.1 | 0.05 | 0.07 |
| Genetic PC11 | -0.11 | 0.06 | 0.05 |
| Genetic PC12 | 0.01 | 0.05 | 0.82 |
| Genetic PC13 | -0.05 | 0.05 | 0.40 |
| Genetic PC14 | -0.02 | 0.05 | 0.67 |
| Genetic PC15 | -0.03 | 0.06 | 0.60 |
| Genetic PC16 | -0.05 | 0.05 | 0.36 |
| Genetic PC17 | -0.03 | 0.05 | 0.64 |
| Genetic PC18 | 0.02 | 0.05 | 0.73 |
| Genetic PC19 | 0.11 | 0.05 | 0.04 |
| Genetic PC20 | 0.03 | 0.05 | 0.58 |
